# Short-term Repeatability of Artificial Intelligence Estimated Electrocardiographic Age

**DOI:** 10.1101/2025.09.04.25335101

**Authors:** Katherine M. Conners, Varun Divi, Elsayed Z. Soliman, Annie Green Howard, Eric A. Whitsel, Didong Li, Michelle Meyer, Christy L. Avery, Faisal F. Syed

## Abstract

Advancements in artificial intelligence have enabled estimation of cardiac age from raw ECG waveforms. ECG-age is a novel metric that provides insights into cardiac function and has shown potential for CVD risk prediction. However, its short-term repeatability remains unclear. ECG-age was measured in 58 participants across two visits, one to two weeks apart. The intraclass correlation coefficient was 0.64 (95% CI: 0.52, 0.76), with a standard error of measurement of 2.6 years. Findings suggest moderate short-term repeatability, with notable within-visit variation. Refining AI algorithms is essential to establish ECG-age as a reliable biomarker for CVD risk assessment and clinical decision-making.

## INTRODUCTION

Contemporary cardiovascular disease (CVD) risk prediction tools rely primarily on vascular indicators, such as blood pressure, cholesterol, and diabetes [1]. Such estimates often lack validation in individuals below the age of 40 and may misclassify risk in certain demographic and clinical subgroups [2]. Assessments of cardiac structure and function offer an attractive alternative to enhance CVD risk stratification, given their ability to predict heart failure, stroke, and sudden cardiac death [3].

Electrocardiograms (ECGs) are essential for CVD diagnosis and provide insights into cardiac structure and function. Recently, artificial intelligence (AI) models have been used to estimate “cardiac age” from ECG waveforms, offering a measure distinct from chronological age, yet correlated. AI-estimated ECG-age reflects the heart’s functional capacity, independent of traditional ECG features, and shows promise in predicting CVD and mortality when significantly higher than chronological age [4]. Despite findings suggesting clinical implications for CVD risk assessment, research on the repeatability of this novel measurement is scarce. Determining its short-term repeatability is crucial for understanding ECG-age and informing its utility in clinical and public health settings.

## METHODS

As detailed previously, 63 participants aged 45-64 years were enrolled at the University of North Carolina at Chapel Hill’s (UNC) General Clinical Research Center between July and October 2011 [5]. Exclusion criteria included pacemaker use, type 1A antiarrhythmic medication, diabetes, congestive heart failure, and renal disease. Participants provided written informed consent, and the study was approved by UNC’s Institutional Review Board. Although distinct from the Atherosclerosis Risk in Communities (ARIC) cohort, this study was conducted under the purview of ARIC personnel, with examinations following their protocols. Participants attended two study visits scheduled one to two weeks apart, after abstaining from caffeine, food, exercise, smoking, and alcohol for 10 hours. During each visit, two 10-second, 12-lead ECG recordings were collected while participants rested in supine position.

We trained and developed a convolutional neural network to predict a person’s ECG-age using the digital waveforms of approximately 24,000 12-lead ECGs from the publicly available German PTB-XL electrocardiography dataset. By directly detecting and extracting features from raw ECG waveforms, the model estimates an individual’s ECG-age independently of traditional ECG features. The model was then applied to UNC participants’ ECG recordings at each of two visits to estimate ECG-age.

We assessed participant-level variability within and between ECG-age measurements as the intra-class correlation coefficient (ICC), calculated as between-participant variance divided by the between + within-participant (total) variance with 95% confidence. We estimated the minimally detectable change (MDC) between the two measurements as follows: MDC95 = SEM×√2_×1.96, where SEM represents the standard error of measurement. The intra-class correlation coefficient (ICC) measures consistency between repeated measurements, while MDC defines the smallest change considered statistically significant. Within-visit differences were defined as the average and absolute differences between ECG-age estimates from the same visit. Between-visit differences were calculated as the average and absolute differences in ECG-age estimates between the two study visits. Lastly, estimated variance was decomposed into between-participant, between-visit, and within-visit components.

## RESULTS

After excluding individuals with ECGs that did not meet quality criteria for our model, data were available for 58 participants. The mean (SD) age was 52 (5) years, 55% were female, and 66% were White. Mean (SD) ECG-age at visits 1 and 2 were 43.5 (4.1) and 44.0 (4.5) years, respectively. The ICC between visits was 0.64 (95% CI: 0.52, 0.76) and the SEM was 2.6 years. The MDC was 7.2 years, representing the smallest statistically significant change beyond measurement error within an individual. The average and absolute within-visit difference in ECG-age were 0.46 years (SD: 3.65) and 3.06 years (SD: 2.00), respectively. For between-visit measurements, the average difference was −0.27 years (SD: 4.10), and the absolute difference was 3.18 years (SD: 2.56). Between-participant variation accounted for 63.8% of the total variance, within-visit variation for 34.6%, and between-visit variation for 1.6% (Table 1).

**Table 1.** Components of Measurement Variation in n=58 University of North Carolina at Chapel Hill’s General Clinical Research Center participants who underwent two ECGs spaced 1-2 weeks apart.

| Source of variation | Standard deviation | % of total variance | Coefficient of variation (%) |
| --- | --- | --- | --- |
| Between-subject | 3.52 | 63.79 | 54.41 |
| Between-visit | 0.56 | 1.64 | 8.72 |
| Within-visit | 2.59 | 34.57 | 40.06 |
| Total | 6.67 | 100.00 | 103.19 |

## CONCLUSION

To our knowledge, this is the first study to assess short-term repeatability of ECG-age. Our findings suggest moderate repeatability over a one-to-two-week interval, indicating that ECG-age remains reasonably stable in the short term. In comparison, ICCs for other ECG parameters range from high (PR interval = 0.93) to lower repeatability (P duration = 0.58), placing ECG-age within the range of traditional ECG metrics. Notable within-visit variation suggests potential influences from measurement error, environmental, or algorithm inconsistencies. Further, the PTB-XL derivation sample included a broader age range and more diverse clinical profiles than our healthy cohort, likely underestimating between-subject variance.

To enhance the clinical applicability of ECG-age, future studies should focus on strategies to reduce within-visit variation, such as refining AI algorithms to minimize technical discrepancies and artifacts. Addressing these issues is essential for establishing ECG-age as a reliable tool for CVD risk assessment. Overall, our findings provide important groundwork for advancing ECG-age as a validated metric in cardiovascular research and clinical workflows.

## Data Availability

All data produced in the present study are available upon reasonable request to the authors

## Acknowledgements

The Atherosclerosis Risk in Communities study has been funded in whole or in part with Federal funds from the National Heart, Lung, and Blood Institute, National Institutes of Health, Department of Health and Human Services, under Contract nos. (75N92022D00001, 75N92022D00002, 75N92022D00003, 75N92022D00004, 75N92022D00005). The authors thank the study participants for their important contribution.

